# Ventilator triggering control with an LSTM-Based Model

**DOI:** 10.64898/2026.04.10.26350573

**Authors:** Jiaying Liu, Jiale Fan, Ziwei Deng, Xuefeng Tang, Hongtao Zhang, Amit Sharma, Qiong Li, Cheng Liang, Yuanyuan Wang, Ling Liu, Kun Luo, Hongde Liu, Haibo Qiu

**Author notes:** **Corresponding author information:** Hongde Liu,; Kun Luo,; Ling Liu,. These authors contributed equally to this work (Jiaying Liu, Jiale Fan, Ziwei Deng).

## Abstract

**Background:** Patient-ventilator synchrony, an essential prerequisite for non-invasive mechanical ventilation, requires an accurate matching of every phase of the respiration between patient and the ventilator.

**Methods:** We developed a long short-term memory (LSTM)-based model that can predict the inspiratory and expiratory time of the patient. This model consisted of two hidden layers, each with eight LSTM units, and was trained using a dataset of approximately 27000 of 500-ms-long flow signals that captured both inspiratory and expiratory events.

**Results:** The LSTM model achieved 97% accuracy and F1 score in the test data, and the average trigger error was less than 2.20%. In the first trial, 10 volunteers were enrolled. In “Compliance” mode, 78.6% of the triggering by the LSTM model was compatible with neuronal respiration, which was higher than Auto-Trak model (74.2%). Auto-Trak model performed marginally better in the modes of pressure support = 5 and 10 cmH_2_O. Considering the success in the first clinical trial, we further tested the models by including five patients with acute respiratory distress syndrome (ARDS). The LSTM model exhibited 60.6% of the triggering in the 33%-box, which is better than 49.0% of Auto-Trak model. And the PVI index of the LSTM model was significantly less than Auto-Trak model (36.5% vs 52.9%).

**Conclusions:** Overall, the LSTM model performed comparable to, or even better than, Auto-Trak model in both latency and PVI index. While other mathematical models have been developed, our model was effectively embedded in the chip to control the triggering of ventilator.

**Trial registration:** Approval Number: 2023ZDSYLL348-P01; Approval Date: 28/09/2023. Clinical Trial Registration Number: ChiCTR2500097446; Registration Date: 19/02/2025.

## 1. Introduction

Ventilators play an important role in both clinic and home in providing respiratory support. Mechanical ventilation is generally categorized into two types modes: passive (controlled) ventilation and active (assisted) ventilation. In passive ventilation, patients are typically deeply sedated or paralyzed, and the ventilator delivers breaths according to preset parameters without any effort from the patient. In contrast, with active ventilation, the patient can initiate or modulate the breathing process themselves, whereby the ventilator must recognize the inspiratory effort and support the whole process accordingly [1].

The coordination between the patient’s respiratory effort and the triggering of mechanical ventilation is extremely important to avoid adverse outcomes in the hospital, especially in “active mode”. This requires a good synchronization between the patient’s neural breathing and the inspiratory and expiratory events triggered by the ventilator [2]. The neuronal respiratory signal can primarily be depicted by the electrical activity of the diaphragm (EAdi). In other words, if a ventilator is triggered via the EAdi signal, synchronization will also be effective[3]. Notably, the setting of the sensing EAdi signal is prone to fall off the skin, causing errors in triggering. Currently, the most common triggering method used in ventilators is the use of airway pressure or flow signals sensed by the ventilator [4]. Chest wall movement and waveform/flow changes in the signal can also be utilized [4]. In the flow or pressure-based method, it must be successfully predicted what type of action the ventilator should resume with inspiratory triggering, expiratory triggering or no triggering. The base model must set a threshold for the flow or pressure, i.e. when the signal reaches the threshold, the ventilator should triggers. In a meta-analysis, the prevalence of ineffective triggering was found to be 0 to 45% when the sensitivity of the flow trigger (cutoff) reached across 0.5 and 6.0 l/min [5]. Furthermore, the occurrence of auto-triggering and ineffective triggering was found to be highly variable and independent of the flow trigger level [5], indicating that more flexible models are warranted.

There has been continuous progress in optimizing different types of signals (such as pressure, flow rate and volume) to trigger ventilators by setting specific cutoffs [6–8]. On this line, Auto-Trak™ system, a model that fully accounts for the escape of air, is now integrated into Philips’ ventilators, has been reported to outperform the conventional flow-cycling model in synchronization, especially in patients with respiratory insufficiency [9]. Recently, Juliette Menguy developed a dynamic prediction model that uses physiological data with machine learning to make more informed decisions about weaning patients from mechanical ventilation, thus reducing the risks associated with prolonged ventilation or premature extubation [10]. Worth mentioning here is IntelliSync+®, an AI-based trigger system that is able to automatically detect transitions between inspiration and expiration by analyzing the morphology of the waveform in real time, making it possible to show improved synchrony, especially in scenarios with significant leakage [11]. Studies have also explored the potential of automatic detection of the patient effort (breathing time) directly from ventilator waveforms using signal processing and machine learning techniques, demonstrating promising accuracy and robustness in detecting inspiratory activity [12, 13].

Patient-ventilator asynchrony (PVA), a frequent and critical problem in mechanical ventilation that leads to clinical problems, is also worth mentioning here. PVA refers to a discrepancy between the patient’s neural breath and the ventilator trigger in terms of time, flow, volume or pressure [14, 15]. PVA can be assessed using several quantitative indicators, including but not limited to triggering delay (latency) and the asynchrony index (AI) or patient-ventilator interaction (PVI) index [16]. Since the EAdi signal is derived from neural breathing, it serves gold standard in assessing the PVA [3]. Both latency and PVI can be estimated from the EAdi signal [17]. Latency represents the percentage of time difference between the ventilator triggering and the patient’s neural breathing. PVI index is a ratio of the number of the PVI events divided to the total respiratory events [18]. PVI events considered mostly include five types asynchrony events, namely early triggering, late triggering, double triggering, failed triggering and artefact. According to the study by Zhou et al., the overall prevalence of PVA in adult critically ill patients over the entire course of mechanical ventilation was 24% [19]. Moreover, the implications and prevalence of the asynchrony have been found to be inconsistent across various studies [20, 21].

Recent studies have leveraged various deep learning architectures to detect PVA, such as recurrent neural networks (RNNs) for real-time identification of missing flow patterns and apneas [22], and convolutional neural networks (CNNs) for the classification of asynchrony types [23].

Although mathematical models have been developed for ventilator triggering, a model embedded in the chip to control the triggering was lack. In this study, we proposed an LSTM-based triggering model specifically designed for passive ventilation, with a particular focus on home-use or non-invasive ventilators. These ventilators are typically employed in chronic care or home settings, where patients may not be able to initiate breath effectively, and sensor resources are limited. Unlike traditional pressure- or flow-threshold triggers, our model uses only flow signals and learns the temporal dynamics of the respiratory cycle to identify inspiratory and expiratory phases in real time. This approach aims to improve synchronization and reduce the incidence of PVA events even in simplified systems with minimal sensor input.

We demonstrated an LSTM-based model that triggers the ventilators based solely on the flow rate signal to accurately predict the patient’s inspiratory and expiratory triggering time. More importantly, an embedded ventilator in the LSTM-based model has been clinically tested.

## 2 Method and dataset

### 2.1 Method

#### The LSTM-based model

The task of the model is to predict which event is needed, inspiratory or expiratory or no triggering, for a patient in the next moment based on the flow rate signal of the last tens of milliseconds. Flow refers to the air velocity detected by ventilator sensor at any given time. Since the flow signal is a kind of time-series data, an LSTM unit was thought to be suitable for modeling.

A simulated medical lung device (IngMar Medical, ASL5000 SW3.5) was used for the model construction data set. (Fig. S1A and Table S1). The input to the model is a numerical vector with a length of 25 points, representing a 500 milliseconds flow rate signal (Fig. S1B). To achieve real-time synchronization with the patient’s breathing rhythm, the input to the model is designed to predict the appropriate ventilator action at each time step. Specifically, the model determines whether an inspiratory phase or an expiratory phase should be triggered or whether the device should remain idle (i.e. not trigger any action). The model is thus formulated as a three-class classification task. The output is a vector of length 3 with values in the range [0,1], corresponding to inspiratory triggering (I), expiratory triggering (E), or no triggering (Non-I/E), respectively (Fig. S1B).

The optimal LSTM model was determined by maximizing the hyperparameters (n=192 hyperparameter combinations). The hyperparameters included number of the hidden layers, number of the LSTM units in each hidden layer, dropout ratio, activation function (call output of each LSTM unit (C and h)), batch size of the inputting data (Fig. S1C and Table 1). We searched for an optimal combination of hyperparameters using a grid search (‘GridSearchCV’ in Python) using five-fold cross-validation algorithm with a loss function of cross-entropy (Eq. 1).

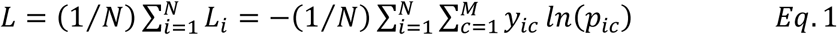

**Table 1.**
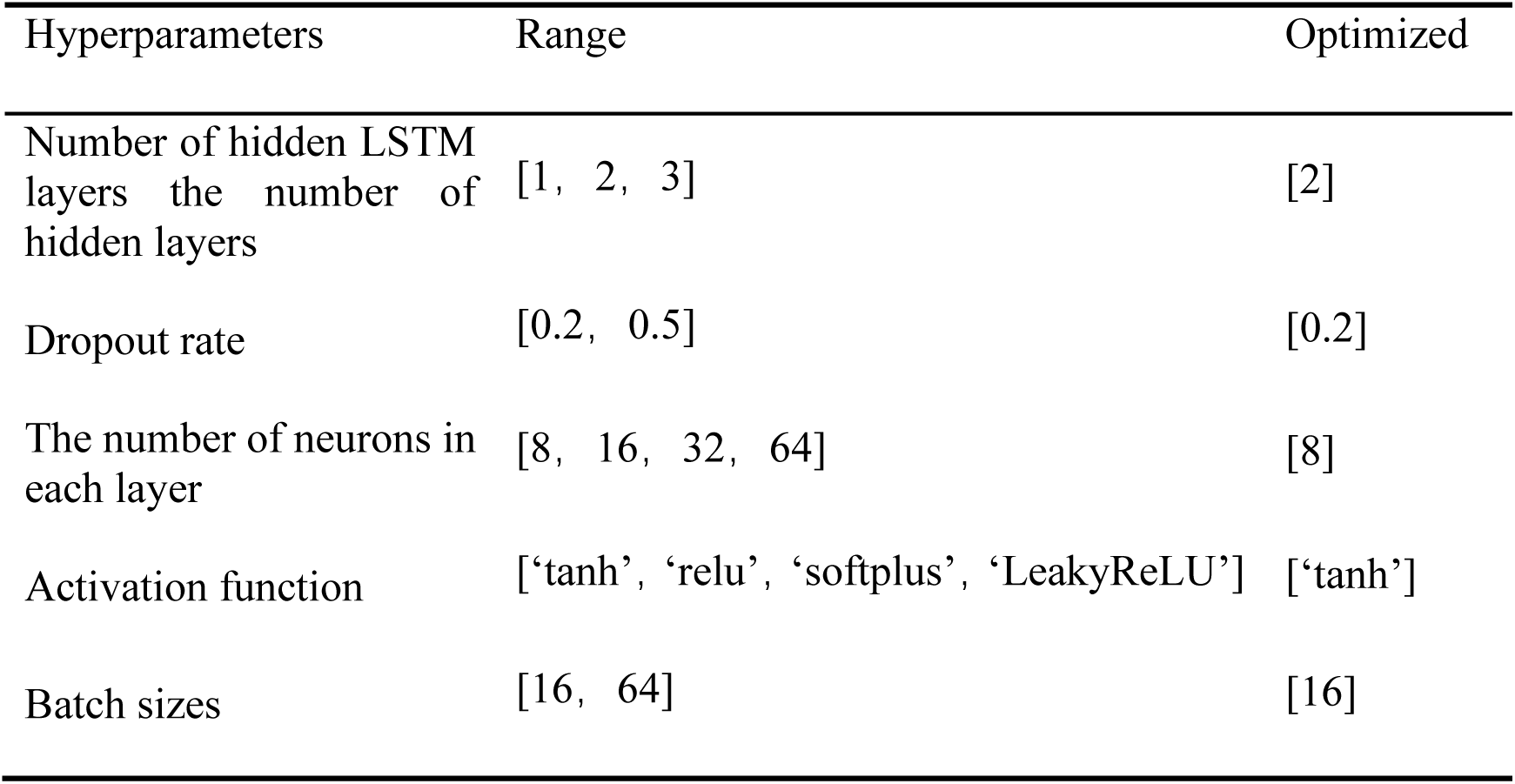
The hyperparameters being optimized and the range of optimization.

| Hyperparameters | Range | Optimized |
| --- | --- | --- |
| Number of hidden LSTM layers the number of hidden layers | [1, 2, 3] | [2] |
| Dropout rate | [0.2, 0.5] | [0.2] |
| The number of neurons in each layer | [8, 16, 32, 64] | [8] |
| Activation function | ['tanh', 'relu', 'softplus', 'LeakyReLU'] | ['tanh'] |
| Batch sizes | [16, 64] | [16] |

Where *N* was the total number of samples and *M* was the number of label categories (I, E, Non-I/E); *y_ic_* was indicator function; and *p_ic_* was the predicted probability of sample *i* belonging to category *c*. The model optimizing algorithm was “adaptive moment estimation” (“Adam” in Python).

Dataset (X, Y) used to construct the model was a three-class data. In data X, each row was a segment of 500 milliseconds flow signal. Each row of Y indicates the class of the corresponding row of X. The performance of the model was evaluated using precision (Eq.2), recall (Eq.3), and F1 score (Eq.4). The error ratio was the proportion of the misclassified samples to the total number of samples for a certain class or multiple classes.

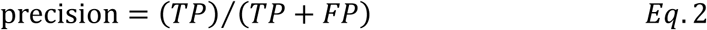

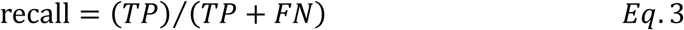

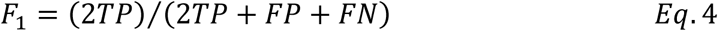

Where *TP*, *FP* and *FN* are true positive, false positive and false negative, respectively. Our model is a three-classification model (I, E and Non-I/E). Given one kind of label, for a signal segment (flow), which corresponds to the label, if the prediction is one of two other labels, it is a FN; if the prediction is the corresponding label, it was a TP. For a segment belongs to other two labels, if the prediction is the label, it is a FP.

The LSTM model was written with Tensorflow (Keras) in Python language. The trained model was then programmed into the ventilator chip (STM32 series) by converting the model (python file) into C language code and then writing it into the chip with STM32Cube.AI tool.

### Clinical trial

We conducted two clinical trials to validate our findings, and for that the research protocol was approved by the Ethics Committee of Zhongda Hospital affiliated to Southeast University of China (Approval No.: 2023ZDSYLL348-P01; Clinical Trial Registration Number: ChiCTR2500097446; Registration Date: 19/02/2025; Approval Date: 28/09/2023). The respiratory system compliance of the volunteers (healthy participants) was assessed using a diaphragm electrode catheter [24]. In the first trial, three types of ventilators, which were controlled with the LSTM model, Auto-Trak model (Philips company), and a cutoff-based model (inspiratory cutoff > 0.5 l/min; expiratory cutoff < 1/3 peak flow), respectively, were compared. Ten volunteers were enrolled in the trial (Table S2). The illustration of the setting for the trial is shown in Fig. S2. EAdi signal was detected with a sensor from company of Maquet Critical Care AB. The sensor had a diameter of 12 Fr. and a length of 125 cm.

The exact procedure of the clinical trial is shown in Fig. S3. The order of ventilator wearing was randomly assigned, and the pressure signal (Paw) of the ventilator and EAdi signal of the sensor were recorded through SerVo Tracker software. The preset ventilator mode was pressure support ventilation (PSV), with the parameters of pressure support (PS) = 5 cmH_2_O, positive end-expiratory pressure (PEEP) = 5 cmH_2_O, and fraction of inspired oxygen (FiO_2_) = 25%. After confirming that the ventilator was functioning normally, it was applied to the subject and data were recorded for 5 minutes. Chest compression was performed to reduce respiratory system compliance, and the level of compression was established before the start of the test. Signals of EAdi and Paw were recorded for 5 minutes. Then, the compression device was removed, and the airway resistance was increased by 20 cmH_2_O, and the EAdi and Paw signals was recorded for 5 minutes. In case that the additional airway resistance returned to 0, the pressure was adjusted to PS = 10 cmH_2_O. Once the breathing curve was stable, the EAdi and Paw signals were recorded for 5 minutes. Following a 5-minute break, another ventilator was connected and the process was repeated.

In the second clinical trial, the LSTM model ventilator was used to support the beathing of five patients with acute respiratory distress syndrome (ARDS). The clinical data of the patients are listed in Table S3. A Philips’ ventilator (Auto-Trak model) was used to compare. The order of ventilator wearing was randomly assigned. EAdi and Paw signals were recorded also with SerVo Tracker software. If the patient showed no intolerance, the wearing time was 5 minutes per treatment. The treatment was cancelled in the event of intolerance.

### Latency of the triggering

The inspiratory and expiratory triggering time was identified in both EAdi and the pressure (Paw) signals (Fig. S4 and Fig. S5A). EAdi signal represents the neural respiratory; and Paw is from the sensor in a ventilator. For EAdi or Paw signals, the identification was done in a sliding window with length of 5 and step of 1. For windows i-1 and i, their difference of mean of the signal was calculated (ΔWmean = Wmeani – Wmeani-1). The standard deviation was represented with Wstdi. the initiation for inspiratory and expiratory was identified through Δwmean and Wstdi. If |ΔWmean| ≤ 2× Wstdi, it is in stationary. Satisfaction of | ΔWmean| > 2× Wstdi means a change in window i, increasing or decreasing. Under this condition, if Wmeani-1 ≤ PEEP, the time is an inspiratory initiation; If Wmeani-1 ≤ (2/3) × PIP, the time is an expiratory initiation (Fig. S4 and Fig. S5A) [25]. Latency of the triggering is defined as in Eq.5 and Eq.6 (Fig. S5B).

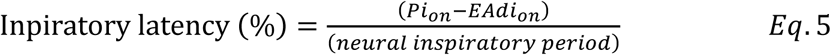

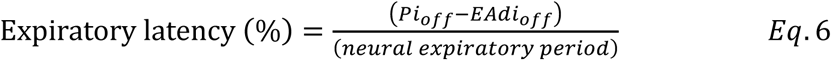

Where *EAdi_on_* and *EAdi_off_* indicated the starting time of neural inspiratory and expiratory on EAdi signal, respectively, while *Pi_on_* and *Pi_off_* indicated the starting time of the inspiratory and expiratory on pressure signal (Paw) ventilator, respectively. Neural inspiratory period in Eq.5 was the time between two neighboring neural expiratory beginnings (*EAdi_off_*), including an inspiratory triggering event (*EAdi_on_*) of the ventilator. If the breath was triggered prematurely, the neural inspiratory period was defined as the interval from the previous *EAdi_off_* to the current *EAdi_on_*. If the triggering was delayed, the neural inspiratory period was defined as the interval from the subsequent *EAdi_off_* to the current *EAdi_on_*. Similarly, neural expiratory period in Eq.6 determined the time between two neighboring neural inspiratory beginnings (*EAdi_on_*), including an expiratory triggering event (*EAdi_off_*). The synchronization was considered acceptable if the both latencies were within ± 33 percent ranges [2].

### Patient-ventilator interaction (PVI) index

PVI index is a ratio of the number of the interaction events divided to the total respiratory events. PVI index indicates the degree of matching of breath phase between patient and the ventilator. Five common PVI events were considered in the test: early triggering, late triggering, failed triggering, double triggering and artefact. Given mean inspiratory time over 30 cycles (I_M_) and inspiratory time (*t*), early triggering refers to *t* ≤ 0.5×I_M_; later triggering, *t* ≥ 2×I_M_; double triggering is defined when two cycles separated by 0.5×I_M_, and the first cycle being patient-triggered. Failed triggering means an abrupt pressure drop (≥ 0.5 cmH_2_O) and flow decrease and not followed by an assisted cycle during the expiratory period. Artefact is defined as a cycle delivered by the ventilator without a prior airway pressure decrease, indicating that the ventilator delivered a breath that was not triggered by the patient [26]. We manually annotated the PVI events by comparing EAdi and Paw signals, under the rules in literature [2, 27].

### 2.2 Datasets

#### Dataset for model construction

For the LSTM model, the training data were collected from the simulated lung medical device produced by IngMar Medical (ASL5000 SW3.5) [28] (Fig. S1A). The device provided flow and pressure signals of 22 populations (Table S1), including starting time of the inspiratory and expiratory events. We extracted the 500 ms-long segments of the flow signal exactly upstream of each inspiratory and expiratory event, and the 500 ms-long segments of non-starting time point. Besides the heathy individuals, we also simulated individuals with different pathologies including ARDS, asthma, pneumonia, chronic obstructive pulmonary disease (COPD). Totally, the total number of segments remained were 27315. Twenty percent of the data (n = 5463) was used as the test set and the remainder was included in the training set. Before training, the amplitude of the flow signal was scaled into range of 0 and 1.

#### Dataset of the clinical trials

In the first clinical trial, we collected data from 300 minutes of EAdi and Paw signals in three modes, compliance, PS = 5 cmH_2_O, and PS = 10 cmH_2_O. The LSTM model, Auto-Trak model and the cutoff-based model accounts for 150, 120 and 30 minutes, respectively (Table S4). In the second clinical trial, we collected 225 minutes of signals of EAdi and Paw signals. The LSTM model, Auto-Trak model and the cutoff-based model accounts for110 and 115 minutes, respectively (Table S5).

## 3 Results

### 3.1 Patterns (shapes) of the flow signal around the inspiratory and expiratory moments

The goal of this work was to trigger a ventilator with an LSTM model using flow signal. The basic idea was that the flow signal changes when an inspiration or expiration event occurs. If the LSTM model is able to adapt with modulated patterns and predict the starting time of the inspiratory and expiratory events in advance, the output of the model can be used to trigger the ventilator.

To test this hypothesis, we first examined the waveform patterns of the flow signal across three classes of data segments: the triggering of inspiration (I), the triggering of expiration (E), and the mid-respiratory phase (Non-I/E). As expected, the averaged flow rate signals before the inspiratory and expiratory events showed the distinct trends (Fig. 1). Around the inspiratory events (I), the flow signal increased (Fig. 1A). Around the expiratory events (E), the flow signal decreased (Fig. 1B). For the mid-respiratory phase (Non-I/E), the flow signal shows a slow change (Fig. 1C). This was consistent with the characteristics of the flow in each phase of breath. We also noticed large variation (variance) in the amplitude of the flow velocity (Fig. 1 A-C), suggesting that the flow varied greatly over time and exhibited multiple kinds of trends, requiring models that could accommodate the diversity of the trends.

**Fig. 1.**
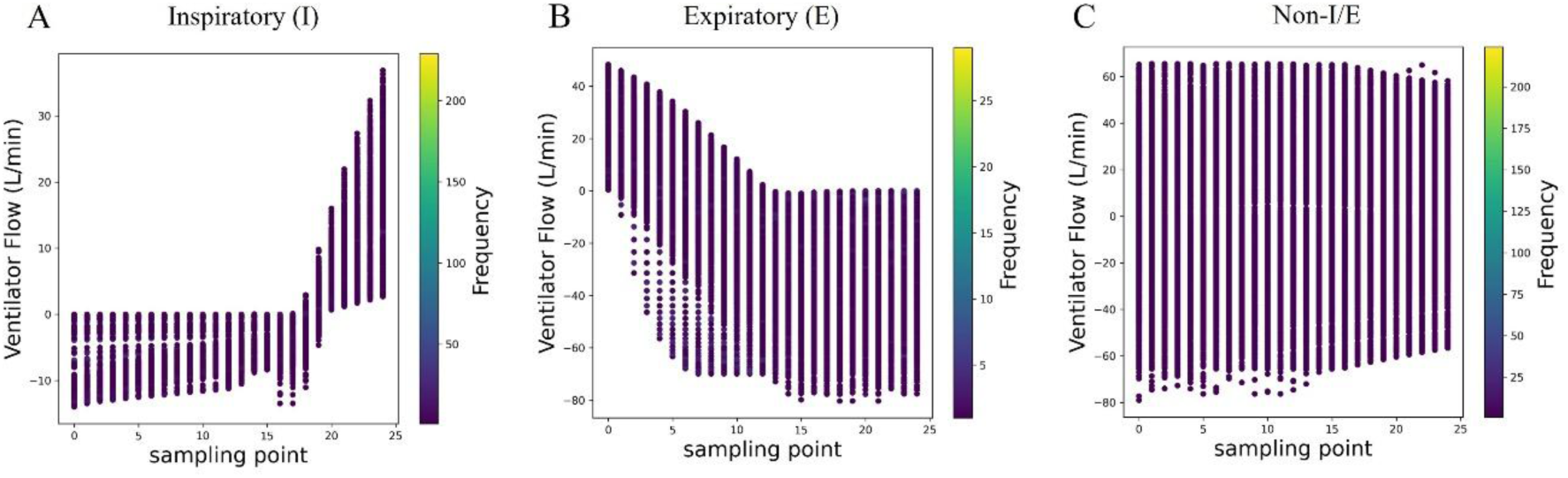
Pattens of flow signals around of the triggering events. A, Averaged signals of flow around the inspiratory (green), expiratory (yellow), and no triggering (blue); Subplots B, C and D are the superposed flow signals for no triggering (Non-I/E), inspiratory (I) and expiratory (E), respectively.

### 3.2 Performance of the optimized LSTM model

In order to establish an optimized network structure, we tested the performance of each network structure by varying the 192 hyperparameters, including number of the hidden layers, number of the LSTM units in each hidden layer, dropout ratio, activation function, batch size of the inputting data (Table 1). In the hyperparameter combinations, 12 network structures showed less in loss function (Eq.1 and Table S6). The models with two hidden layers and with activation function “tanh” appeared to be better (Table S6). In model selection, another factor we considered was the size of the network. Since we needed to integrate the trained model into ventilator chip, the smaller size was considered to be optimal. Finally, a network with two hidden layers and each layer had eight LSTM units activated with function “tanh” (2Layers-8Cells, tanh) was selected (Table 1 and Fig. 2A).

**Fig. 2.**
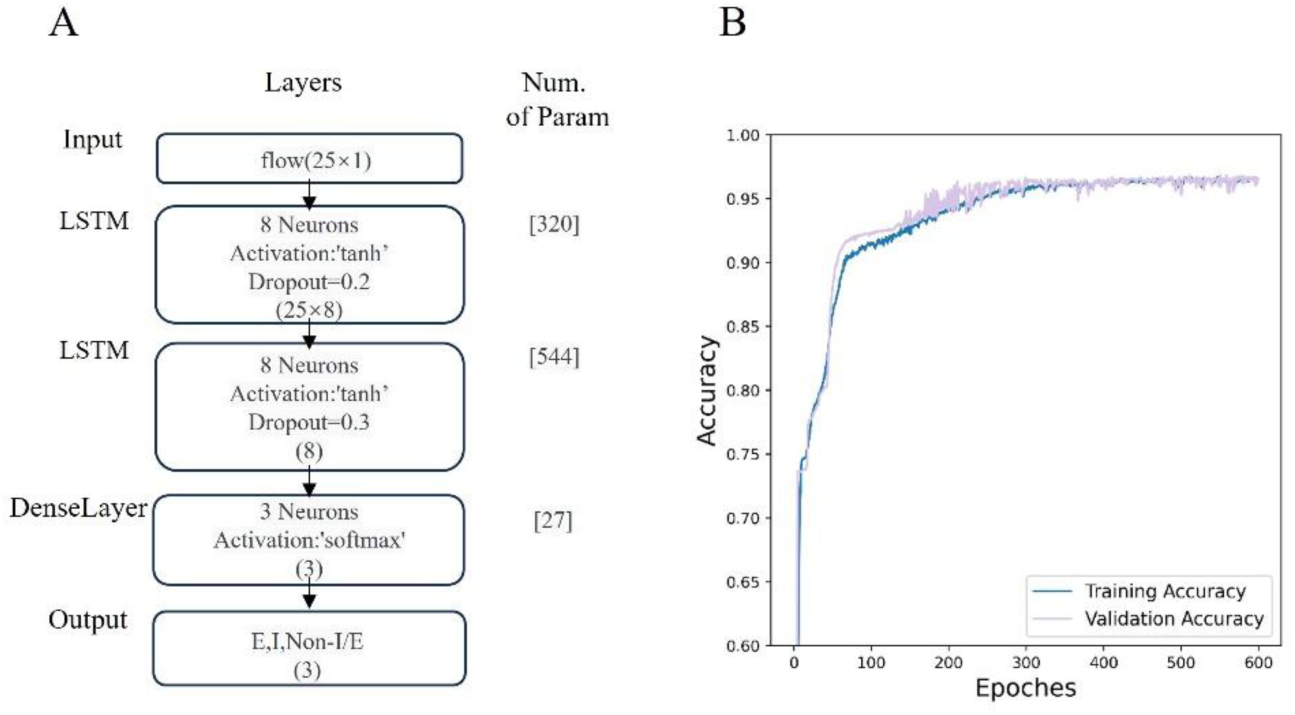
The optimized LSTM model and its performance in training. A, Structure of the optimized LSTM model. The model consists of two LSTM layer and a dense layer; B and C, the loss (B) And the accuracy (C) In training and validation processes over 600 epochs;

In training with optimizer ‘Adam’ in Python, the optimized LSTM model converged quickly after 600 epochs (Fig. 2B). The loss value dropped significantly at the beginning of learning, indicating that the model’s structure and the optimization algorithm were appropriate. When the model learned to a certain extent, the loss value tended to be stable and found to be fluctuating around 5% (Fig. 2B). The accuracy increased correspondingly to the decrease of loss value in the training (Fig. 2B).

On the test data (20% of the 27315 samples), the prediction error ratio for three kinds of labels (I, E and Non-I/E,) were 0.038, 0.037, and 0.01, (Tables 2 and 3), respectively, indicating a good performance. The prediction error for the Non-I/E labels was lower than two other two labels. It was probably the pattern of Non-I/E was relatively monotonous and could be easily captured by the model. The lower error was helpful in reducing double triggers and artifact events, as the models of ventilation control frequently encounters non-I/E situations in real-world applications.

**Table 2.** Confusion matrix of the prediction by the LSTM model on the test data.

| Real Labels | Predicted by the LSTM model |  |  |
| --- | --- | --- | --- |
|  | Non-I/E | Expiratory(E) | Inspiratory(I) |
| Non-I/E | 3057 | 15 | 16 |
| Expiratory(E) | 35 | 1142 | 9 |
| Inspiratory(I) | 37 | 8 | 1144 |

**Table 3.** Performance of the optimized LSTM model on the test data. Weighted average is called using weight of the support number to total number for three classes.

|  | Precision | Recall | F1-Score | Error ratio | Support |
| --- | --- | --- | --- | --- | --- |
| Non-E/I | 0.98 | 0.99 | 0.98 | 0.010 | 3088 |
| E | 0.98 | 0.96 | 0.97 | 0.037 | 1186 |
| I | 0.98 | 0.96 | 0.97 | 0.038 | 1189 |
| Weighted Average | 0.98 | 0.98 | 0.98 | 0.022 | 5463 |

**Table 4.** Patient-ventilator interaction (PVI) index (%) in the first clinical trial for ten volunteers.

| Ventilator<br>PVI events | The cutoff-based model |  |  | Auto-Trak |  |  | The LSTM model |  |  |
| --- | --- | --- | --- | --- | --- | --- | --- | --- | --- |
|  | PS5 | PS10 | C | PS5 | PS10 | C | PS5 | PS10 | C |
| Early triggering | 7.8 | 38.2 | 50.1 | 7.1 | 4.3 | 8.4 | 9.4 | 5.6 | 4.5 |
| Late triggering | 32.2 | 10.2 | 1.3 | 20.5 | 11.8 | 12.1 | 21.1 | 20.2 | 26.2 |
| Failed triggering | 9.3 | 0.0 | 0.0 | 1.8 | 0.7 | 0 | 0.2 | 0.0 | 0.0 |
| Double triggering | 0.0 | 11.5 | 4.8 | 0.2 | 0.0 | 0.0 | 0.0 | 0.4 | 0.0 |
| Artefact | 0.6 | 1.2 | 2.4 | 2.6 | 20.6 | 10.7 | 6.5 | 3.5 | 4.3 |
| Total | 49.9 | 61.1 | 58.6 | 32.2 | 37.4 | 31.2 | 37.3 | 29.7 | 35 |

Note: Modes: PS5, PS = 5 cmH<sub>2</sub>O; PS10, PS = 10 cmH<sub>2</sub>O; C, Compliance.

The weighted averaged error ratio of prediction was 2.20% (Table 3). Precision, recall, F1 score and the error rate were listed in Table 3. It was acknowledged that the error rate in practice was below 5%. This indicates that the performance was adequate.

### 3.3 Evaluations in the clinical trial

We then tested three types of ventilators, which were triggered with the LSTM model, Auto-Trak, and a cutoff model, respectively. The test was carried out in three modes, “PS = 5 cmH_2_O”, “PS = 10 cmH_2_O” and “compliance”. Mode “PS = 5 cmH_2_O” provided a lower level of support and was suitable for patients with mild dyspnea or those who needed less assistance. Mode “PS = 10 cmH_2_O” provided a higher level of support and was suitable for patients with moderate dyspnea or those who needed more assistance. Mode “compliance” referred to the patient’s ability to trigger expiration and expiration without much effort.

In the first clinical trial, respiratory data were collected from 10 volunteers over a total duration of 300 minutes. The LSTM model accounted for 150 minutes, Auto-Trak model accounted for 120 minutes and the cutoff model accounted for 30 minutes (Table S4).

The recorded EAdi was used as the standard to evaluate the performance of the ventilators’ triggering. The triggering time and the neural staring time of inspiratory and expiratory were identified with a decision algorithm (Fig. S4). The latencies of inspiration and expiration were calculated according to the formula in Fig. S5B (Eq.5 and Eq.6) [2]. Taking *EAdi_on_* as a reference, if the pressure triggering (*Pi_on_*) started earlier than *EAdi_on_*, it was considered early; if it started later, it was considered late.

The latencies for the three models in three modes of the testing were shown in Fig. 3A, where the dashed boxes indicated that triggering latencies of both inspiratory and expiratory within 33% of the neural breath defined by EAdi signal. Latency in the 33%-box means an accepted matching between the neural respiratory and the triggering of the ventilators. For the mode “Compliance”, 78.6% of the triggering by the LSTM model were in the box, indicating a well matched to the neural respiration, which was slightly higher than Auto-Trak model (74.2%). In mode “PS = 10 cmH_2_O”, Auto-Trak model performed best (94.6%) (Fig. 3A), followed by the LSTM model (92.7%). In mode “PS = 5 cmH_2_O”, the Auto-Trak was better than the LSTM model (88.5% vs 78.8%). Although the percentages were distinct, the statistical significance was not obvious between the LSTM model and Auto-Trak model (Chi-square test, Compliance: p = 0.93; PS = 5 cmH_2_O, p = 0.37; PS = 10 cmH_2_O, p = 0.69) (Table S7). This suggested that the performance was comparable between the two models in triggering latency. In three modes, the cutoff model-based ventilator performed the worst. Also, the results indicated that a higher-pressure support led a less latency and a better matching. Even for the cutoff-based model, in mode “PS = 10 cmH_2_O”, there was 82.4% of the triggering in the 33%-box (Fig. 3A).

**Fig. 3.**
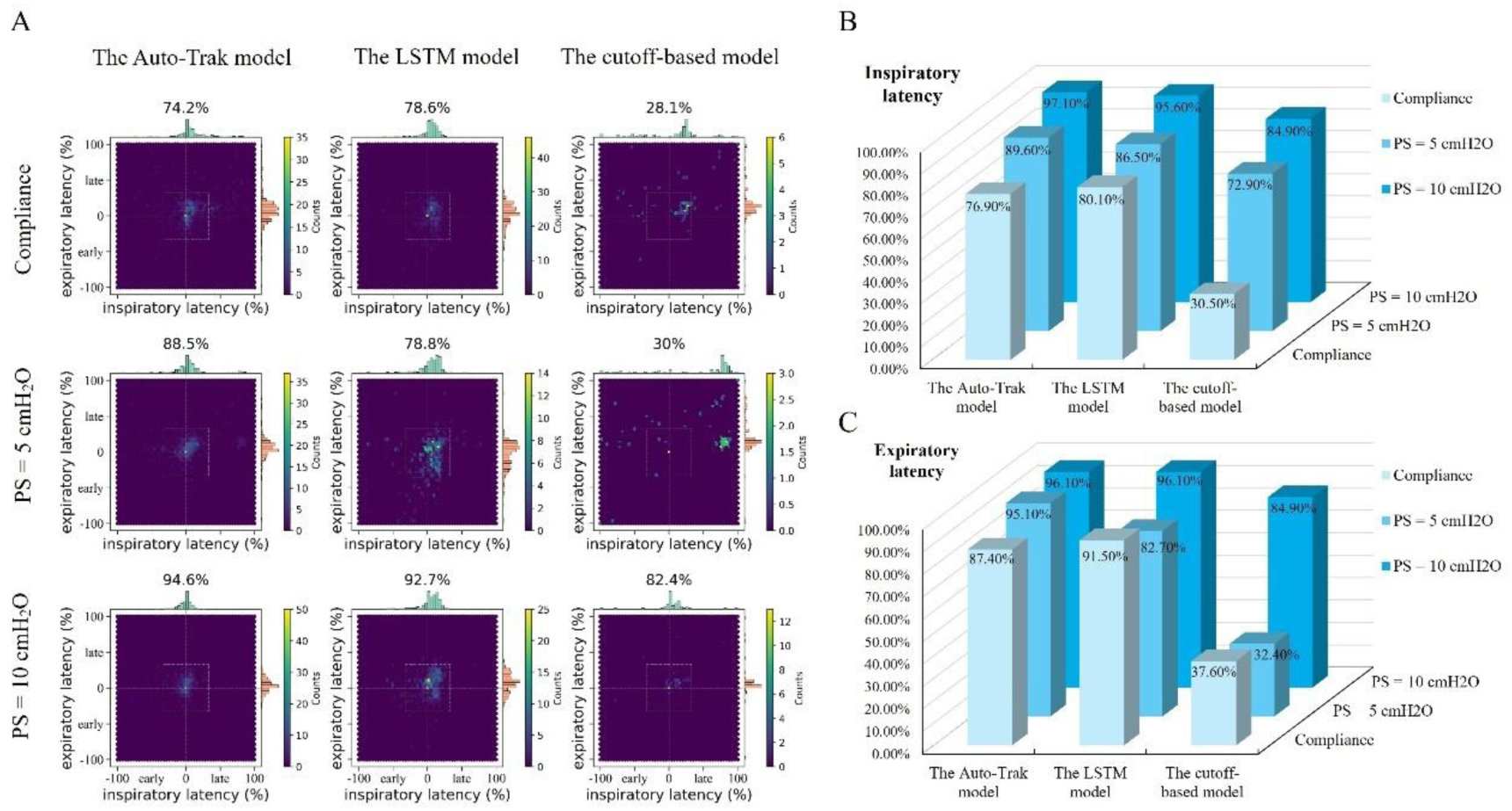
Latencies of triggering in the three models using the electrical activity of the diaphragm (EAdi) as a criterion in the first clinical trial. A, The latencies of the inspiratory and expiratory triggering events by the three models (the LSTM model, Auto-Trak, and a cutoff-based model) in three testing modes (compliance, PS = 5 cmH_2_O, and PS = 10 cmH_2_O). Dotted box in each panel indicated region of the latency less than 33% in comparing to the neural respiration. The percentage of the triggering events less than 33% of latency is indicated on the top of each panel. B, The percentage of the latency falling into 33%-box for the inspiratory and expiratory, respectively.

We also calculated the percentage of the latency falling into 33%-box for the inspiratory and expiratory phases (Fig. 3B and C), respectively. The results indicated that in “Compliance” mode, the LSTM model was better than the Auto-Trak model. In “PS = 10 cmH_2_O” mode, the LSTM model was very similar to the Auto-Trak in both inspiratory and expiratory triggering. We noticed that in the “PS = 5 cmH_2_O” mode, the LSTM model had a delayed latency compared to Auto-Trak model in expiratory triggering.

To further validate the model performance, the PVI index was calculated (Tables S8 and S9. The index varied greatly among the volunteers, probably due to the response difference of the volunteers. Notably, Auto-Trak and LSTM models exhibited a low PVI. In mode “Compliance”, the PVI index of Auto-Trak model was 31.2%, while the LSTM model was 35.0% (Table S8). Auto-Trak model was slightly better than our model. In mode “PS = 10 cmH_2_O”, the LSTM model had a significant lower PVI index (29.7%) than Auto-Trak model (37.4%) (p = 0.0029) (Table S9). In mode “PS = 5 cmH_2_O”, both showed a very close PVI index (32.2% vs 37.3%) (p = 0.034) (Table S9). A lower PVI index means a better synchrony. This resulted indicated that the LSTM was comparable and even better the Auto-Trak.

In the second clinical trial, the LSTM model demonstrated better performance compared to the Auto-Trak model) (Fig. 4). Specifically, in terms of triggering latency, 60.6% of the triggering by the LSTM model was in the 33%-box (Fig. 4A), indicating a well match with the neural respiration. For the Auto-Trak, only 49.0% of the triggering was in that box (Fig. 4B). Significance of the difference was p = 0.088 (Table S7). Moreover, the LSTM model showed a significant lower PVI index than the Auto-Trak (36.5% vs 52.9%, p = 2.3×10^−15^) (Tables 5, S10 and S11). For five PVI events, the LSTM model had a significant lower PVI index than the Auto-Trak, except double triggering (Table 5). Lower PVI index means a better synchronization. The LSTM model was superior to the Auto-Trak model in the clinical trial for the ARDS patients.

**Fig. 4.**
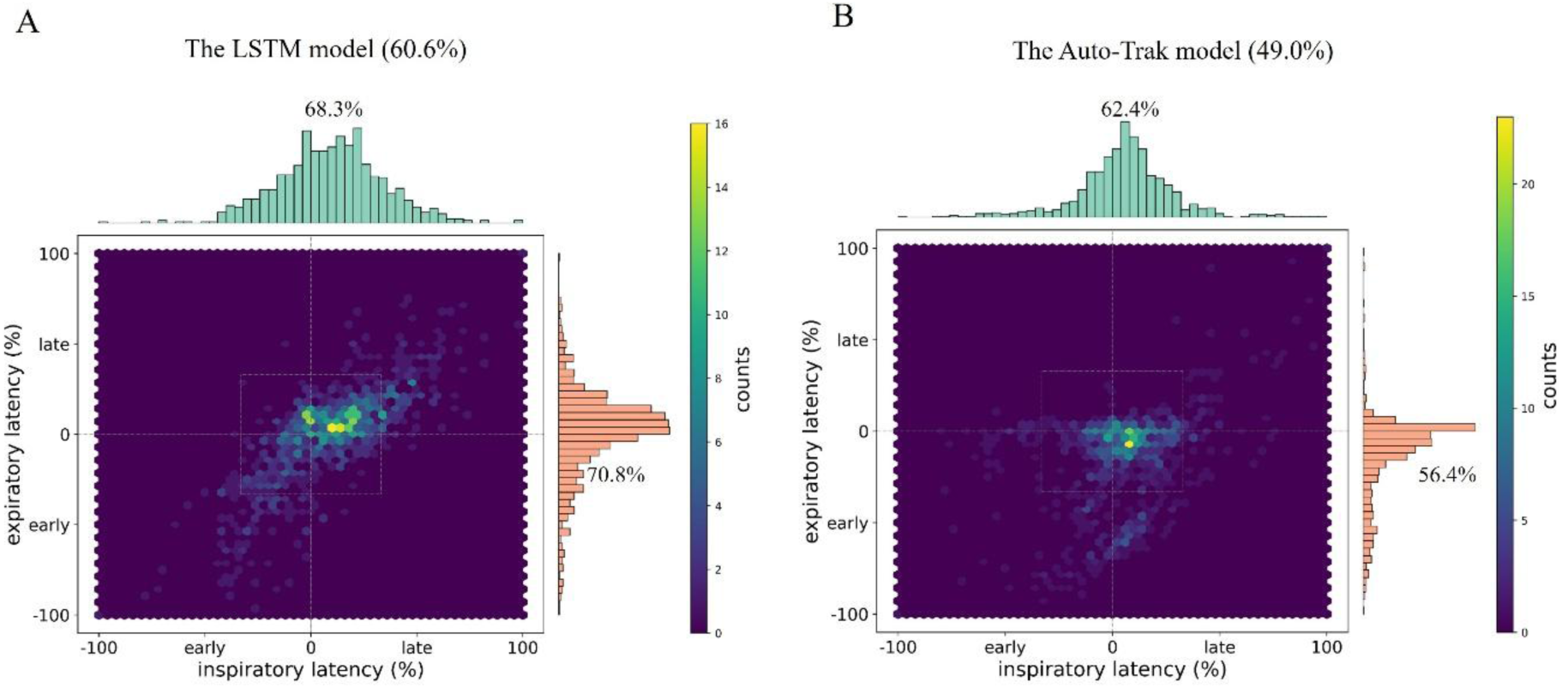
Latencies of triggering for the ARDS Patients. The latencies of the inspiratory and expiratory triggering events by the LSTM model (A) and the Auto-Trak model (B). Dotted box in each panel indicated region of the latency less than 33% in comparing to the neural respiration. The percentage of the triggering events less than 33% of latency is indicated on the top of each panel. The percentages for the inspiratory and expiratory triggering are indicated at top and right of each panel, respectively.

**Table 5.**
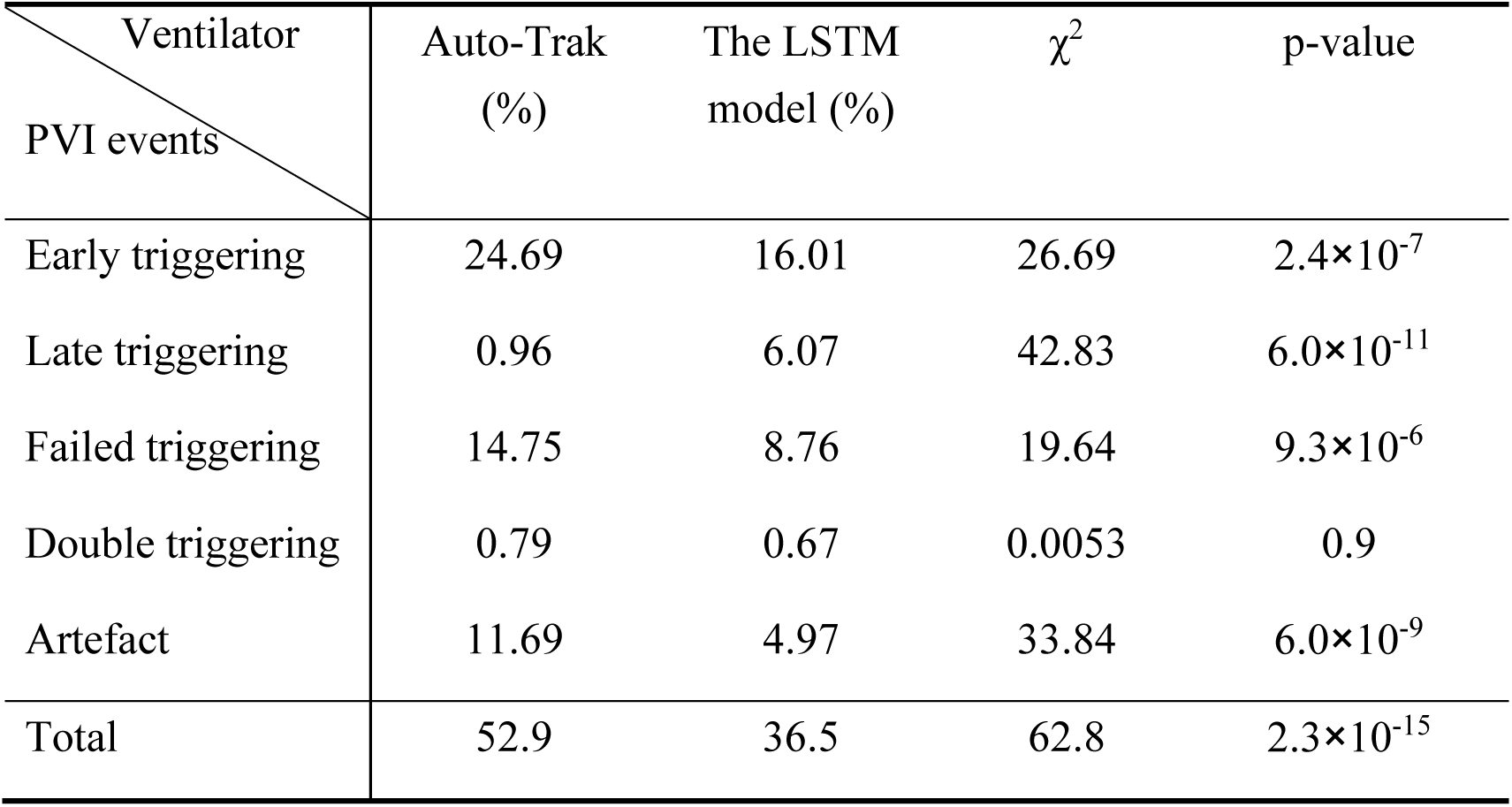
Patient-ventilator interaction (PVI) index (%) in the second clinical trial (For five ARDS patients).

## 4 Discussion

Despite mechanical ventilation being an extremely important part of clinical care, asynchrony between patient and ventilator can occur if the ventilator is not synchronized properly with the patient’s breathing cycle. Therefore, the automatic synchronization between human and ventilator is the most important performance indicator of a ventilator. Therefore, automatic human-ventilator synchronization is the most critical performance indicator of a ventilator. In particular, optimal respiratory triggering control should be synchronized with neural autonomic respiration [29]. Here, we have developed an LSTM-based model primarily using the flow rate signals. When controlling the triggering with the LSTM model, two key points were considered: (1) the synchronization should be accurate, and (2) the model should be lightweight enough to operate on microcontroller platforms for embedded applications.

In the clinical trial for the heathy volunteers, the LSTM model showed comparable performance to the Auto-Trak model. Of importance, the LSTM model showed shorter latencies and lower PVI indices in ARDS patients, better than the Auto-Trak model While other mathematical models have been developed for ventilator triggering, our model was effectively embedded in the chip to control the triggering. We therefore offer an alternative, AI-based option for controlling a ventilator.

Nevertheless, some limitations in our methodology should be acknowledged. First, the dataset used to construct the model was generated from a simulated lung device. This likely does not capture the waveform complexity of real-world population, especially patient with the lung pathology. Second, the LSTM model did not account for air leakage, which may have lower stability predictions. Third, our model relies exclusively on the flow signal and lack information about air pressure. Finally, the total number of the study participants in the trial is limited and a large-scale clinical trial is needed.

In summary, our LSTM-based triggering model shows promise as an AI-driven solution for human-ventilator synchronization, especially in resource-limited or home-care settings. With further development and validation, it may serve as an effective alternative or complement to current triggering mechanisms.

## 5 Conclusion

An LSTM model was generated to trigger the ventilators on the basis of flow signals. In clinical performance tests, the LSTM model was comparable or even better to Auto-Trak model in terms of synchronization and latency. We demonstrated the feasibility of controlling ventilators with a neural network model.

## Declarations

### Abbreviations

PVA: Patient ventilator asynchrony
PEEP: Positive end of expiratory pressure
PVI: Patient ventilator interaction
LSTM: Long short-term memory
RNN: Recurrent neural network
PS: Pressure support
FiO2: Fraction of Inspired oxygen
PSV: Pressure support ventilation
PAW: Pressure of Airway
EAdi: Electrical activity of the diaphragm

### Ethics approval and consent to participate

All ethics of the current research have been approved by the Ethics Committee of Zhongda Hospital affiliated to Southeast University of China (Approval No.: 2023ZDSYLL348-P01; Clinical Trial Registration Number: ChiCTR2500097446; Registration Date: 19/02/2025; Approval Date: 28/09/2023). All patients have signed and submitted their consent to participate in the research and their data privacy has been fully considered. Informed consent was obtained from all subjects. All methods were carried out in accordance with relevant guidelines and regulations (Declaration of Helsinki).

### Consent for publication

Not applicable.

### Availability of data and materials

Access to data is restricted due to security and privacy considerations. However, data supporting the findings of this study are available upon reasonable request.

### Competing interests

The authors declare no competing interests.

### Funding

The work was supported by “Tianshan Talent” Training Program (2023TSYCLJ0030), National Key Research and Development Program of China (2022YFC2504405), and Jiangsu Provincial Medical Key Laboratory (ZDXYS202205).

### Authors’ contributions

DHL, KL and YYW conceived the study and trained the model. LL, JLF, ZWD and JYL carried out the clinical trial and data analysis. HDL, AS, QL, and JLF wrote the manuscript. XFT, HTZ, HBQ and ZLS provided critical feedback and discussion.

## Data Availability

All data produced in the present study are available upon reasonable request to the authors.

## Acknowledgements

The sponsor had no role in the design of the study, the collection and analysis of the data, or the preparation of the manuscript.

## Supplementary material

### Tables

**Table S1 Dataset for training and testing model.**

**Table S2 Information about ten volunteers in the first clinical trial.**

**Table S3 Information about five patients in the second clinical trial.**

**Table S4 Test duration in three modes (Compliance, Pressure support (PS) = 5 cmH_2_O, and PS = 10 cmH_2_O) for each ventilator in the first clinical trial (minutes).**

**Table S5 Test duration in the second clinical trial for each ventilator (minutes).**

**Table S6 Loss on the test data for the combinations of number of hidden layers, number of cells (LSTM units) in each layer and activation function types.**

**Table S7 Significance the latency percentage in 33%-box between the SLTM model and Auto-Trak.**

**Table S8 Patient-ventilator interaction (PVI) events in the clinical trial for ten volunteers.**

**Table S9. Chi-squared test of PVI index between Auto-Trak and the LSTM model.**

**Table S10 PVI events for the five patients with the LSTM model ventilator, PVI index was indicated in the last row**

**Table S11 PVI events for the five patients with the Auto-Trak model ventilator, PVI index was indicated in the last row.**

### Figures

**Figure S1 Model construction dataset generation and the hyperparameters in the LSTM model frame.** A, GUI of the simulated lung device (ASL5000 SW 3.5, InMar Medical); B, The sampling procedure for three-class flow segments (Inspiratory (I), Expiratory (E), and non-triggering (Non-I/E)); C, The LSTM model frame and the hyperparameters to optimize.

**Figure S2 Demonstration of the setting in the clinical trial.** A, Electrical activity of the diaphragm (EAdi) signal was sampled by placing a sensor into the esophagus. EAdi signal was recorded with SerVo Tracker device. The sensor was from Maquet Critical Care AB. Volunteer or patient wears a ventilator. Pressure signal (pressure of airway (PAW)) in the ventilator was also recorded with SerVo Tracker device. B, The sensor of Maquet Critical Care AB, and with a diameter of 12 Fr and a length of 125 cm.

**Fig. S3 The procedure of the clinical trial.**

**Fig. S4 The algorithm of identifying the initiation time for inspiratory and expiratory from Eadi and Paw signals.** For Eadi or Paw signals, the identification was done in a sliding window with length of 5 and step of 1. The difference in the signal mean value was calculated for windows i-1 and i, their difference of mean of the signal was calculated (Δwmean = Wmeani – Wmeani-1). The standard deviation is represented with Wstdi. The initiation for inspiratory and expiratory was identified through Δwmean and Wstdi. If |Δwmean| ≤ 2× Wstdi, it is in stationary. Satisfaction of | Δwmean| > 2× Wstdi means a change in window i, increasing or decreasing. Under this condition, if Wmeani-1 ≤ PEEP, the time represents an inspiratory initiation; If Wmeani-1 ≤ (2/3) × PIP, the time reflects an expiratory initiation.

**Fig. S5 A, Identification of the starting time for inspiration and expiration on Eadi and airway pressure (Paw) signals.** *EAdi_off_* and *EAdi_on_* indicated the beginning of neural expiration and inspiration, respectively; and *Pi_off_* and *Pi_on_* indicated the triggering of inspiration and expiration, respectively. Shown was for the random segments; B, Demonstration of the latency calculation; neural inspiratory and expiratory periods as defined according the EAdi signals. Difference between the starting time of neural respiration and the triggering time by the models was calculated and scaled with the neural periods. A small latency means a good synchronization.

## Notes

**Funding information:** The work was supported by “Tianshan Talent” Training Program (2023TSYCLJ0030), National Key Research and Development Program of China (2022YFC2504405), and Jiangsu Provincial Medical Key Laboratory (ZDXYS202205).

### Competing Interest Statement

The authors have declared no competing interest.

### Clinical Trial

Clinical Trial Registration Number: ChiCTR2500097446

### Funding Statement

The study was funded by Tianshan Talent Training Program (2023TSYCLJ0030), National Key Research and Development Program of China (2022YFC2504405), and Jiangsu Provincial Medical Key Laboratory (ZDXYS202205).

### Author Declarations

Ethics committee/IRB of Zhongda Hospital affiliated to Southeast University gave ethical approval for this work

